# SUDMEX CONN: The Mexican MRI dataset of patients with cocaine use disorder

**DOI:** 10.1101/2021.09.03.21263048

**Authors:** Diego Angeles-Valdez, Jalil Rasgado-Toledo, Victor Issa-Garcia, Thania Balducci, Viviana Villicaña, Alely Valencia, Jorge Julio Gonzalez Olvera, Ernesto Reyes-Zamorano, Eduardo A. Garza-Villarreal

**Affiliations:** Instituto de Neurobiología, Universidad Nacional Autónoma de México campus Juriquilla, Querétaro, Mexico; Escuela de Medicina y Ciencias de la Salud TecSalud, Tecnológico de Monterrey, Monterrey, Mexico; University of Groningen, Department of Biomedical Sciences of Cells and Systems, Cognitive Neuroscience Center, University Medical Center Groningen, Groningen, the Netherlands; División de estudios de posgrado de la Facultad de Medicina, Universidad Nacional Autónoma de México, Mexico; Université de Bordeaux, Bordeaux, France; Subdirección de Investigaciones Clínicas, Instituto Nacional de Psiquiatría Ramón de la Fuente Muñız, Mexico City, Mexico; Comisión Nacional para la Prevención de Adicciones, Mexico City, Mexico; Faculty of Psychology, Universidad Anáhuac México Sur, Mexico City, Mexico

## Abstract

Cocaine use disorder (CUD) is a substance use disorder (SUD) characterized by compulsion to seek, use and abuse of cocaine, with severe health and economic consequences for the patients, their families and society. Due to the lack of successful treatments and high relapse rate, more research is needed to understand this and other SUD. Here, we present the SUDMEX CONN dataset, a Mexican open dataset of CUD patients and matched healthy controls that includes demographic, cognitive, clinical, and magnetic resonance imaging (MRI) data. MRI data includes: 1) structural (T1-weighted), 2) multishell high-angular resolution diffusion-weighted (DWI-HARDI) and 3) functional (resting state fMRI) sequences. The repository contains unprocessed MRI data available in brain imaging data structure (BIDS) format with corresponding metadata available at the OpenNeuro data sharing platform. Researchers can pursue brain variability between these groups or use a single group for a larger population sample.

## Background and Summary

Substance use disorders (SUD) are considered a worldwide health problem characterized by patterns of continuous psychoactive substance seeking, use and abuse despite consequences ^1^. These substances include alcohol, cannabis, nicotine, opioids, and stimulants such as cocaine ^2^. Cocaine use disorder (CUD) is described as the compulsive searching behavior for cocaine consumption which produces several alterations and plasticity changes in reward processing and impulsiveness. CUD is also accompanied with cognitive function deficits in executive functions and verbal memory during withdrawal that could be reversible depending on use frequency and abstinence period ^3–5^.

The complexity of CUD behavior has led to research with several techniques including neuroimaging. These techniques arise the opportunity to identify biomarkers and in diagnostic processes, providing information about neurobiological effects of substance use that could be linked with subjective experiences and behavior, and allows the comparison of brain structure and function with healthy controls ^6,7^.

The use of non-invasive neuroimaging techniques, such as magnetic resonance imaging (MRI), have helped to elucidate some of the neurobiological mechanisms and alterations in SUD. In a recent meta-analysis on SUDs, studies reported lower gray matter and white matter volume in the thalamus, claustrum, insula, inferior frontal gyrus, and superior temporal gyrus in individuals with CUD ^8^. Similarly,^9^ studies have also reported consistent pathology in diffusion MRI studies in the genu of the corpus callosum, with possible implications in interhemispheric communication and cognitive functions. Alterations aforementioned areas have been shown to undergo functional connectivity changes that compromise processes such as reward signal, executive function, and impulsivity ^10^. Taken together, studies using MRI techniques could help to understand CUD pathology and contribute to the improvement of therapeutic approaches.

Our dataset comes from a cross-sectional case-control study with the goal of understanding the clinical, cognitive, neuroanatomical and functional pathology that crack cocaine induces. It includes a sample of 75 (9 female) CUD patients and 62 (11 female) healthy controls (HC). The acquired MRI sequences comprise whole-brain: 1) structural (T1-weighted), 2) novel multishell high angular resolution diffusion-weighted (DWI-HARDI) and 3) functional (resting state fMRI) sequences. Participants also underwent comprehensive clinical and cognitive evaluations. A participant checklist with the specific MRI sequence acquired can be found in Supplementary 2. Four features from this dataset can be highlighted: 1) the use of the same scanner, protocol and operating procedure, 2) the CUD group shows greater movement, commonly found in psychiatric samples, 3) the novel multi-shell DWI protocol which allows for further study of brain structure and 4) the extensive clinical and cognitive evaluations the participants underwent.

Previous publications have used this dataset using diffusion kurtosis imaging analysis ^11^, neurite orientation dispersion and density imaging analysis ^12^, and a machine learning approach for the identification of cognitive markers in CUD ^13^, among others ^,14–17^. Overall, this dataset could contribute to the in-depth study of substance use disorders, particularly cocaine use.

## Method

### Participants

We scanned a total of 146 participants (patients and controls). From the total sample, nine participants were eliminated (Supplementary 1 and 3). Thus, the final sample consisted of 75 CUD participants (nine female) and 62 HC (six female) scanned from March 2015 to October 2016. Demographic characteristics ^18^ of the sample are summarized in Table 1. All participants provided verbal and written informed consent. The study was carried out according to the Declaration of Helsinki and was approved by the Ethics Committee of the Instituto Nacional de Psiquiatría “Ramón de la Fuente Muñiz”. Cocaine dependence was diagnosed in CUD patients using MINI International Neuropsychiatric Interview - Plus Spanish version 5.0.0 ^19^ which was administered by trained psychiatrists. The participants were also assessed with clinical and cognitive tests.

**Table 1.** Demographic characteristics of dataset

|  | HC<br>(n = 62) | CUD<br>(n = 75) | Statistic |
| --- | --- | --- | --- |
| Age | 30.6 ± 8.26 | 31 ± 7.20 | $t(122) = -0.30$<br>$p = 0.76$ |
| Sex |  |  |  |
| Male | 51 (82.25%) | 66 (88%) | $\chi^2(1) = 0.49$ |
| Female | 11 (17.71%) | 9 (12%) | $p = 0.48$ |
| Years of consumption | n.a | 10.1 ± 6.8 | n.a |
| AMAI score | 133.44 ± 50.6 | 107.29 ± 50.6 | $t(118) = 2.29$<br>$p < 0.05$ |
| Degree |  |  |  |
| Elementary | 3 (4.83%) | 6 (8%) | $\chi^2(5) = 14.05$ |
| Middle school | 14 (22.58%) | 32 (42.66%) | $p = 0.015$ |
| High school | 16 (25.80%) | 10 (13.33%) |  |
| Technical | 5 (8.06%) | 12 (16%) |  |
| Bachelor degree | 21 (33.87%) | 13 (17.33%) |  |
| Doctorate degree | 1 (1.61%) | 0 (0%) |  |

Data are given as N(%) or mean (SD), Two samples *t*-test was performed for age, and $\chi^2$ for categorical variables, n.a: no applica. HC, healthy control; CUD, cocaine use disorder.

To be considered for the study, CUD patients had to be active cocaine consumers or with an abstinence period shorter than 60 days prior to the scan; frequency of use had to be at least 3 days per week with no more than 60 continued days of abstinence during the last 12 months. Additional exclusion criteria for all groups were: medical, neurological and other psychiatric disorders, severe suicidal risk, history of head trauma with loss of consciousness, and non-compliance with magnetic resonance imaging safety standards. Recruitment criteria are shown in Supplementary 1.

### Magnetic Resonance Imaging Acquisition

MRI sequences were acquired using a Philips Ingenia 3 T MR system (Philips Healthcare, Best, The Netherlands, and Boston, MA, USA), with a 32-channel dS Head coil. The order of the sequences was the following for the single session: 1) resting state (rs-fMRI), 2) T1-weighted (T1w) and 3) High Angular Resolution Diffusion Imaging (DWI-HARDI). This order was maintained across participants. Previous to the MRI acquisition, the amount of alcohol in participants blood was measured using a breath alcohol test. Total scan time was approximately 50 min. During the study the participants were fitted with MRI-compatible headphones and goggles.

### Functional Images and field maps

Resting state fMRI sequences were acquired using a gradient recalled (GE) echo planar imaging (EPI) sequence with the following parameters: dummies = 5, repetition time (TR)/echo time (TE)=2000/30.001ms, flip angle=75°, matrix=80×80, field of view=240mm^2^, voxel size=3 × 3 × 3mm, slice acquisition order=interleaved (ascending), number of slices=36, phase encoding direction=AP. The total scan time of the rs-fMRI session was 10 min with a total of 300 volumes acquired. All participants were instructed to keep their eyes open, to relax while not thinking about anything in particular. We used MRI-compatible goggles (fiber optic glasses SV-7021, Avotec) to show the participants a fixation cross (white cross with black background) and we used the included eye-tracking camera to prevent participants falling asleep during this sequence. If the participants closed their eyes for more than 10 seconds, we would wake them up using the communication through the headphones, reminding them to try to not fall asleep for the 10 minutes the sequence lasted and we re-started the sequence over.

To create field maps intended for rs-fMRI use, we acquired opposite phase sequences, using the same GE-EPI sequence with the following parameters: TR/TE=2000/30ms, flip angle=75°, matrix=80×80, voxel size=3×3×3mm, number of slices=36, phase encoding direction=PA. A total of 4 volumes were acquired.

### Anatomical and diffusion images

T1w were acquired using a three-dimensional FFE SENSE sequence, TR/TE=7/3.5 ms, field of view=240mm^2^, matrix=240 × 240 mm, number of slices=180, gap=0, plane=sagittal, voxel=1 × 1 × 1 mm (first five participants were acquired with a voxel size=0.75 × 0.75 × 1 mm). DWI-HARDI used a SE sequence, TR/TE =8600/126.78 ms, field of view=224mm^2^, matrix=112 × 112 mm, number of slices=50, gap=0, plane=axial, voxel=2 × 2 × 2 mm, directions: 8 = b0, 36 = b-value 1,000 s/mm2 and 92 = b-value 3,000 s/mm2, total = 136 directions.

Field maps intended for DWI-HARDI use were acquired using a spin echo (SE) EPI sequence with the following parameters: TR/TE=8600/127ms, flip angle=90°, matrix=112×112, voxel size=2×2×2mm, number of slices=50, phase encoding direction=PA. A total of 7 volumes were acquired.

## Data Records

### Format Organization

The dataset follows the Brain Imaging Data Structure (BIDS, v. 1.0.1) organization and is available and updated on the OpenNeuro Data sharing platform (https://Openneuro.org). BIDS organization are used to facilitate data sharing and unify the majority of projects in the field by using simple folder structure, standardized file names accordingly to acquisition modality in NIfTI format (converted from DICOM) with data descriptions and metadata in JavaScript Object Notation (JSON) files ^20^. DICOM to NIfTI conversion was performed using dcm2bids v. 2.1.4, which uses dcm2niix v. 1.0.201 ^21^. All subjects were anonymized using pydeface to remove facial features ^22^. Please find further details in the corresponding documentation (https://bids.neuroimaging.io/).

### Quality Control

Quality assessment of the MRI dataset was run using image Quality Measures (MRIQC v. 0.15: an automatic prediction of quality with visual reports of each sequence) from structural T1w and rs-fMRI images, available in OpenNeuro platform. This tool helps to extract and compute different metrics such as signal-to-noise ratio, a noise model fitting parameters, and spatial distribution of information. All of these can be exported to HTML and JSON reports ^23^. Please find further details in corresponding documentation (mriqc.readthedocs.io/).

### Clinical measures

Participants were evaluated with a set of paper-based clinical tests before MRI scanning. The tests included were: 1) Mini International Neuropsychiatric Interview - Plus, 2) Addiction Severity Index, 3) Structured Clinical Interview for DSM-IV Axis II Personality Disorders, 4) Symptom Checklist-90-Revised, 5) Cocaine Craving Questionnaire General and Now, 6) World Health Organization Disability Assessment Schedule, 7) Barratt Impulsiveness Scale, 8) Edinburgh Handedness Inventory Short Form, 9) Difficulties in Emotion Regulation Scale, 10) Clinical Global Impressions Scale, 11) Revised Diagnostic Interview for Borderlines, 12) Dissociative Experiences Scale, 13) 4-item Dissociation-Tension Scale and 14) Instant view. These clinical tests were carried out by trained mental health psychologists and psychiatrists, in a quiet room without distractions:

#### Mini International Neuropsychiatric Interview - Plus (M.I.N.I.- Plus)

The M.I.N.I. is a comprehensive diagnostic psychiatric interview designed to assess the 17 most common psychiatric disorders of the Axis I in DSM-IV and the ICD-10 ^24^. We used the official Spanish translation version 5.0. of the M.I.N.I.-Plus, an extended version that includes a total of 23 psychiatric disorders. The interview consists of a set of structured questions that require a yes or no answer. These questions are designed to cover all the diagnostic criteria of the different psychiatric disorders. The interview is divided into modules that correspond to the diagnostic category which was applied by qualified psychiatrists.

#### Addiction Severity Index (ASI)

The ASI is a semi-structured interview designed to assess the lifetime and the past-30-days status in several functional domains: alcohol and drug use, medical and psychiatric health, employment, self-support, family relations, and illegal activities ^25^. We used the Spanish translation ^26^ of the revised 5th edition ^27^.

#### Structured Clinical Interview for DSM-IV Axis II Personality Disorders (SCID-II)

The SCID-II is a psychiatric interview that allows the diagnosis of DSM-IV personality disorders ^28^. The instrument has a self-administered questionnaire appended in which DSM-IV Axis-II criteria are asked in the form of yes/no questions. We only applied this SCID-II questionnaire. It first characterizes briefly the typical behavior, relationships and capacity for self-reflection of the interviewee and then assesses each of the personality disorders. We used the official Spanish translation ^29^.

#### Symptom Checklist-90-Revised (SCL-90-R)

The SCL-90-R is a self-report measure of psychological symptoms and distress ^30^. It includes 9 symptom dimensions (somatization, obsessive-compulsive, interpersonal sensitivity, depression, anxiety, hostility, phobic anxiety, paranoid ideation, and psychoticism) and 3 global indices of distress: Global Severity Index (GSI), Positive Symptom Distress Index (PSDI) and Positive Symptoms Total (PST). To score the instrument, an average of all the items for each symptom dimension is obtained. The GSI is calculated by calculating a global average of the 90 items. The PSDI is the amount of non-zero responses given by the participant, while PST is calculated by the sum of the 90 items’ scores divided by the PSDI. In general, for the 9 symptom dimensions and the 3 global indices of distress, values above the 90th percentile are considered high, indicating a person at risk. We used a Spanish translation^31^.

#### Cocaine Craving Questionnaire General (CCQ-General) and Now (CCQ-Now)

The CCQ is a 45-item questionnaire that explores cocaine craving among patients ^32^. The CCQ-General asks participants to rate their craving over the previous week. The CCQ-Now asks participants about their craving at the moment of assessment. Both versions of the CCQ consist of the same items, written in different tenses. The items are related to the following contents: desire to use cocaine, intention and planning to use cocaine, anticipation of positive outcome, the anticipation of relief from withdrawal or dysphoria, and lack of control overuse. We used the CCQ-General Spanish translation that was validated in Mexican population ^33^.

#### World Health Organization Disability Assessment Schedule 2.0 (WHODAS 2.0)

The WHODAS is an instrument designed to evaluate functional impairments or disabilities of patients with psychiatric disorders ^34^. The WHODAS 2.0 is an updated version of the instrument ^35^. We used the official 36-item, self-administered, Spanish translation of this version ^36^.

#### Barratt Impulsiveness Scale Version 11 (BIS-11)

The BIS was developed more than 50 years ago ^37^. The BIS has been extensively revised into the BIS-11 ^38^. This version of the scale consists of 30 items that describe impulsive and non-impulsive behaviors related to 3 main categories: attentional, motor, and non-planning impulsiveness. We interpreted the total scores as suggested in a later publication ^39^ : highly impulsive (total score ≥ 72), within normal limits for impulsiveness (total score 52-71), extremely over-controlled or has not honestly completed the questionnaire (total score < 52). We used the Spanish translation of the BIS-11 ^40^.

#### Edinburgh Handedness Inventory - Short Form (EHI-SF)

The Edinburgh Handedness Inventory (EHI) is used for objectively determining whether someone is right or left-handed ^41^. We used the Short Form of this inventory to assess handedness on all the participants, as there is evidence supporting an enhancement in handedness classification ^42^. The EHI-SF asks participants to rate their hand preference when performing 4 activities (4 selected items out of the 10 from the original inventory): writing, throwing, toothbrush, and spoon. The responses to the 4 items are then averaged to calculate a laterality quotient, from which handedness can be classified (left handers = −100 to −61, mixed handers = −60 to 60, right handers = 61 to 100). We translated the EHI-SF into Spanish for use in this study.

#### Difficulties in Emotion Regulation Scale (DERS)

The DERS is a scale with 24 items and five subscales: non-acceptance of emotional responses, difficulty engaging in goal-directed behavior, lack of emotional awareness and lack of emotional clarity ^43^. We used the validated Spanish translation of scale ^44^.

#### Clinical Global Impressions Scale (CGI) for Borderline Personality Disorder (CGI-BPD)

The CGI is a brief, stand-alone clinical scale that evaluates a wide variety of psychiatric disorders based on clinician’s view of the patient’s global functioning. The CGI modification evaluates specifically BPD status. It is commonly utilized to measure the disorder severity and improvement by a given intervention, including global improvement and index of efficacy ^45,46^.

#### Revised Diagnostic Interview for Borderlines (DIB-R)

The DIB-R is a semi-structured interview used for a standardized diagnosis and clinical severity of BPD. The measures that provide are the major aspects of the disorder: affect, cognition, impulse action patterns and interpersonal relationships ^47^. We used the Spanish translation of instrument ^48^.

#### Dissociative Experiences Scale (DES)

The DES is a self-report scale that allows the clinicians to quantify dissociative experiences. The scale considers five areas of dissociative disorders described in DSM-IV. As a screening instrument makes it possible to differentiate between participants with clinical diagnosis of a dissociative disorder and those without ^49^. We used the Spanish translation of instrument ^50^.

#### 4-item Dissociation-Tension Scale (DSS-4)

The DSS-4 is a short self-rating instrument for assessment of dissociative symptoms including psychological and somatic aspects. The instrument is sensitive for intervention treatments and is commonly used in neuropsychological and neuroimaging experiments ^51^. We translated DSS-4 into Spanish for use in this study.

#### Instant-View™ Multi Drug Urine Test

We performed Instant-View™ Multi Drug Urine Test (catalog number 03-3635) from Kabla™ (Monterrey, NL, Mexico) on participants prior to performing the MRI study. The screening test uses lateral flow chromatographic immunoassay technique with reagent strips to detect the presence of drugs above a cutoff (the following in the catalog number we used; amphetamine: 1000 ng/mL, benzodiazepines: 300 ng/mL, cocaine: 300 ng/mL, methamphetamine: 1000 ng/mL, morphine/opiates: 2000 ng/mL, marihuana: 50 ng/mL) and/or its metabolites in urine. The test was performed following the manufacturer instructions.

### Cognitive measures

For measuring cognitive domains, participants were administered several cognitive tests in two different modalities: computer-based using the Psychology Experiment Building Language (PEBL) version 2.0 with Spanish translation ^52^, and paper-based. All tests administered via PEBL were: 1) Berg’s Card Sorting Test, 2) Flanker Task, 3) Go/No-go task, 4) Letter-number 5) Digit span backward, 6) Iowa gambling task, 7) Tower of London and 8) Reading the Mind in the Eyes Test. The Digit-Span Backward and Letter-Numbers Sequencing were paper-based tests. A trained psychologist administered the cognitive tests in a quiet room to avoid distractions. The duration of the tests was approximately 45 minutes for each participant, and it was performed after the MRI scan.

#### Cognitive flexibility

Berg’s Card Sorting Test (BCST): The computer version of the Wisconsin Card Sorting Test consists of a 128-card deck. Each card is presented individually to the participants to discover the rule and match the given card with key cards shown on the screen by shape, color, or quantity. Once ten cards are matched correctly, the sorting rule changes ^53,54^.

#### Inhibition

Flanker task: The task measures response inhibition processes by the identification of stimuli over noise. Participants observed a rapidly changing row of arrows and must respond to the middle arrow and ignore the others in a target-distracting sequence ^55,56^.

Go/No-go task: In this task, participants must respond quickly to a signal stimulus (go) and withhold the response with a no-go signal. The ability to inhibit the response during no-go signals is measured along with response time (RT) ^57,58^.

#### Working memory

Letter-number sequencing: A series of letters and numbers are presented to participants and are required to remember and repeat each series sequentially, starting with numbers and followed by the letters in ascending order and alphabetical ^59^.

Digit span backward: A list of numbers presented by the examiner must be repeated in reverse order by the participants. Difficulty increases every two items and stops if participants fail two times in a row ^59^.

#### Decision making

Iowa gambling task (IGT): Participants observe four virtual decks of cards. They turn over one at a time from any deck until 100 trials. Then, they receive or lose a certain amount of money according to a previously determined advantageous or disadvantageous pattern. Participants start with $2000 and the objective is to maximize profits ^60,61^.

#### Planning

Tower of London (ToLo): This task is extensively used to measure planning ability and other executive functions. Participants observe a top board as a goal model and must preplan a sequence of moves one token at a time to reproduce it on the bottom board with the lowest movements possible ^62,63^.

#### Theory of mind

Reading the mind in the eyes test (RMET): The objective of this task is to detect others’ emotional states, feelings and thinking. Participants observe 36 photographs of grayscale around the upper face, identify it and select one of four options on the screen to match the photograph with the mental state word. To reach this task, participants need to have a mental state lexicon and know the presented terms ^64^.

## Technical Validation

The quality of MRI sequences was evaluated using the MRIQC v. 0.15 assessment. For high-resolution structural images (T1-weighted) here we show: 1) mean cortical thickness (Fig. 1), 2) Signal-to-Noise Ratio (SNR) (Fig. 2A), 3) contrast-to-noise ratio (Fig. 2B) and 4) entropy-focus criterion (Fig. 2C). For rs-fMRI images we extracted the following metrics: 1) mean framewise displacement (Fig. 2D), 2) temporal Signal-to-Noise Ratio (Fig. 2E), and 3) spatial standard deviation of successive difference images (Fig. 2F). For diffusion images, we extracted the SNR for corpus callosum (Fig. 3).

**Figure 1.**
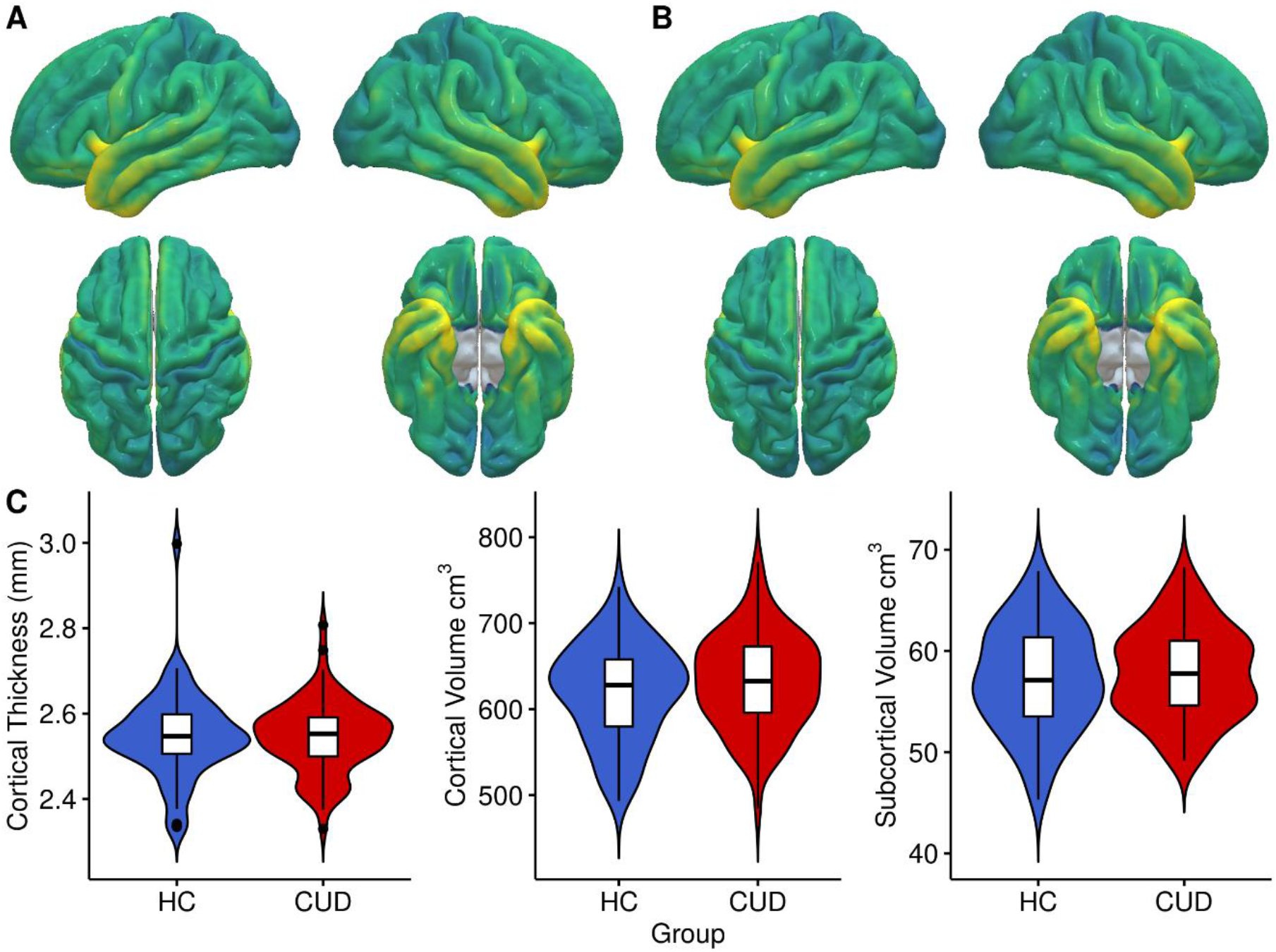
Anatomical MRI metrics for each group. Mean cortical thickness of (A) Healthy Control (HC), and (B) Cocaine Use Disorder (CUD) group. (C) Cortical Thickness (mm) and Volume (cm^3^) extracted for each group.

**Figure 2.**
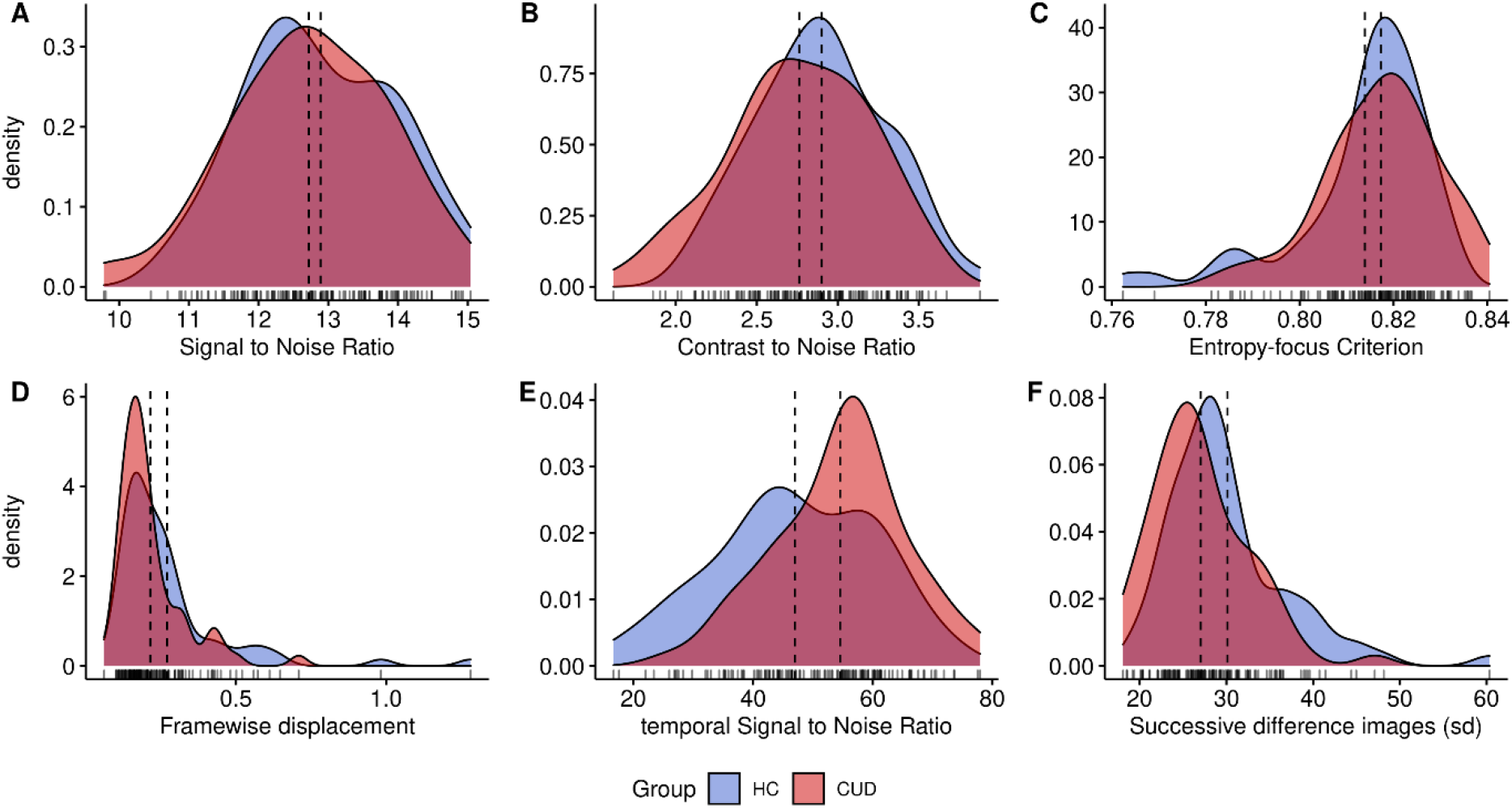
Quality metrics for anatomical and functional images. Anatomical images: (A) Signal-to-Noise Ratio (SNR), (B) contrast-to-noise ratio, (C) entropy-focus criterion. Functional images: (D) mean framewise displacement, (E) temporal Signal-to-Noise Ratio, and (F) spatial standard deviation of successive difference images, for healthy control (HC) and cocaine use disorder (CUD).

**Figure 3.**
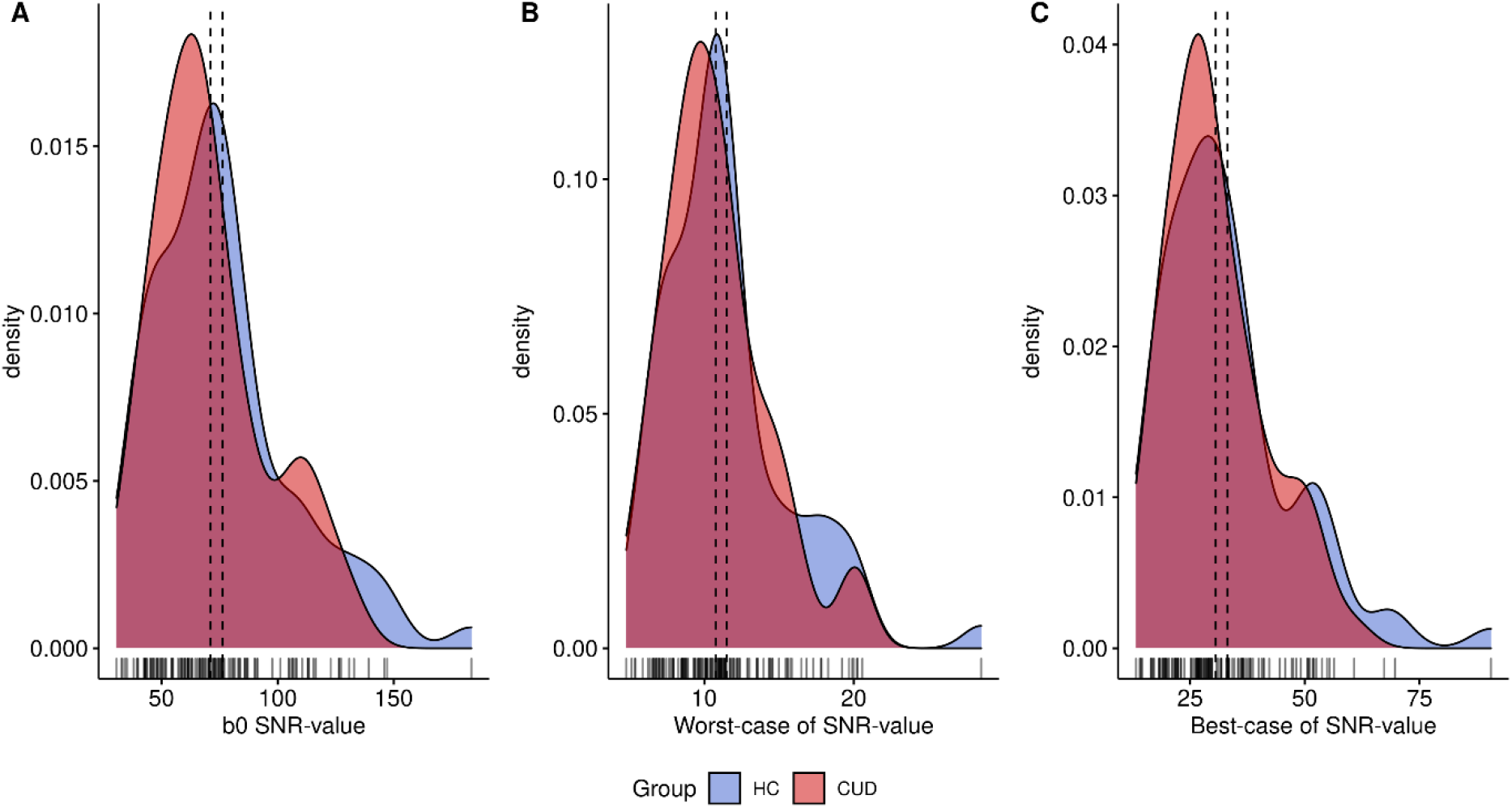
Quality metrics for diffusion images. Signal-to-Noise Ratio values (SNR) for Diffusion-weighted images computed in corpus callosum for (A) B0, (B) Worst-case SNR value and (C) Best-case, for healthy control (HC) and cocaine use disorder (CUD).

Global mean cortical thickness (Fig. 1A-B) was calculated using FreeSurfer v.7 ^65^ for each group, which is a measure of the cortical gray matter width over the surface of the brain based on T1-weighted images ^66^. In the same way, volumes (Fig. 1C) were obtained as the count of all voxels that are in total of cortical gray matter and in subcortical gray matter, these metrics are commonly associated with alteration according to demographic or genetic variables and in several pathologies such as CUD ^67^.

Signal in MRI refers to the mean voxel intensity in the image, in contrast to the random differences in voxel intensity which is considered as noise ^68,69^. The ratio of the mean signal intensity and noise is defined as Signal-to-Noise ratio. SNR was extracted from MRIQC output for T1w and rs-fMRI images. Moreover, for DWI, SNR within the Corpus Callosum was calculated using Diffusion Imaging in Python (Dipy; https://dipy.org/documentation/) protocol estimation ^70^. We extracted the worst (Fig. 3B) and best case (Fig. 3C) of SNR besides the b0 (Fig. 3A).

Contrast-to-noise ratio (CSN), extracted for structural T1w images, is an extension of SNR for quantitative noise measure, with the advantage of not being influenced by contrast or brightness changes, where higher values indicate better quality ^71^. Entropy-focus criterion (EFC), also extracted for structural images, is a ghosting and blurring indicator through the Shannon entropy measure ^72^. Framewise displacement (FD), used for rs-fMRI images, is described as the sum of translational and rotational realignment parameters of instantaneous head motion ^73^. Spatial standard deviation of successive difference images (DVARS) were calculated to estimate noise variance of rs-fMRI signals across the brain ^74^.

## Data Availability

The MRI data is available for download at https://openneuro.org/datasets/ds003346. Please download the latest available version as there may be updates. Clinical and cognitive measures are available in Zenodo http://doi.org/10.5281/zenodo.5123331.

http://doi.org/10.5281/zenodo.5123331

## Usage Notes

The present MRI dataset is composed from patients with cocaine use disorder and healthy controls. Uses include scientific research and academic purposes. Researchers can use novel analysis, compare variance within techniques, for the pursuit of brain variability between these groups or use one of them for a larger population sample. All clinical and cognitive data are available in Zenodo and accompany each participant’s MRI data. We recommend the use of framewise displacement (FD) and artifacts correction methods for preprocessing data due to high motion during scanning.

## Acknowledgments

We thank the people who helped this project in one way or another: Alan Dávalos, Adedamola Falana, Francisco J. Pellicer Graham, Margarita López-Titla, Aline Leduc, Erik Morelos-Santana, Lya Paas, Daniela Casillas, Sarael Alcauter, Luis Concha and Bernd Foerster. We also thank Rocio Estrada Ordoñez and Isabel Lizarindari Espinosa Luna at the Unidad de Atención Toxicológica Xochimilco for all their help and effort. Finally, we thank the study participants for their cooperation and patience. Victor Issa-Garcia and Eduardo A. Garza-Villarreal would like to thank Dirección General de Calidad y Educación en Salud, Secretaría de Salud, México for the scholarship support provided to Victor. This project was funded by CONACYT-FOSISS project No. 0201493 and CONACYT-Cátedras project No. 2358948.

## Author contributions

E.G.V., and J.J.G.O. originated the concept of dataset. D.A.V., T.B., and A.V. recruited and acquired the MRI data. T.B, V.V. and E.R.Z. developed and implemented the cognitive and clinical battery. D.A.V., J.R.T., and V.I.G, performed the analysis and technical validation D.A.V., J.R.T., V.I.G. and E.G.V. created and edited the manuscript.

## Competing interests

The authors declare no conflict of interest.

## Notes

### Competing Interest Statement

The authors have declared no competing interest.

### Funding Statement

This project was funded by CONACYT-FOSISS project No. 0201493 and CONACYT-Catedras project No. 2358948.

### Author Declarations

The study was carried out according to the Declaration of Helsinki and was approved by the Ethics Committee of the Instituto Nacional de Psiquiatria Ramon de la Fuente Muniz.

