## Supplementary Material 1 for "SUDMEX CONN: The Mexican MRI dataset of patients with cocaine use disorder"

For the manuscript:

**Supplementary 1. Study criteria.**

**Inclusion**

- Minimum age of 18 years and maximum of 50 years old.
- Cocaine use for at least 1 year, with current average use of at least 3 times a week, with periods of continuous abstinence of less than one month during the last year.
- Desire to participate and agree to the informed consent.

**Exclusion**

- First-degree personal or family history of any clinically defined neurological disorder.
- Any electronic or metal implants or device (i.e., aneurysm clips, shunts, stimulators, cochlear implants, or electrodes).
- Splinters of metal or metal projectiles to the head or body.
- Current use of any investigational drug or of any medicine with anti- or pro-convulsive action such as tricyclic antidepressants or neuroleptics, unless prescribed for craving symptoms.
- History of schizophrenia, bipolar disorder, mania, or hypomania.
- History of any heart condition currently under medical care (i.e., myocardial infarction, angina pectoris, congestive heart failure, etc.)
- Women with reproductive potential not using an acceptable form of contraception, as well as pregnant or lactating women.
- Current dependence (by DSM-IV criteria) on substances other than cocaine and / or nicotine (cocaine use disorder).
- Claustrophobia.

**Elimination**

- Expressed desire to stop participating.
- Those who presented abnormal radiological findings warranting clinical attention outside the study to ensure the health of the participant.
- The appearance of psychotic symptoms related to addictive disorder.
