## Supplementary Material 3 for "SUDMEX CONN: The Mexican MRI dataset of patients with cocaine use disorder"

For the manuscript:

**Supplementary 3. Eliminated participants**

**RID	Motive**

39	Diabetes Mellitus

63	No MRI

69	Diabetes Mellitus

79	No MR

89	No MRI

90	No MRI

96	No MRI

102	Diabetes Mellitus / hypertension

113	No MRI

119	Hypertension

121	No MRI

135	No MRI

143	No MRI

148	No MRI

150	No MRI

151	No MRI

152	No MRI

155	No MRI

157	No MRI

158	No MRI
